# The Rapid Occluded MCA Vessel Etiology (ROME) Score - Identifying the Etiology of Large Vessel Occlusions of the Middle Cerebral Artery

**DOI:** 10.1101/2025.03.26.25324735

**Authors:** Michael Fana, Omar Choudhury, Katie Latack, Lonni Schultz, Abdallah Albanna, Taylor Reardon, Zahid Iqbal, Max Kole, Horia Marin, Alex B. Chebl

## Abstract

**Background:** Differentiating between intra-cerebral atherosclerotic disease (ICAD) and non-ICAD large vessel occlusion (LVO) is crucial for selecting the appropriate mechanical thrombectomy (MT) technique and device. We developed an algorithm to predict LVO etiology using clinical and radiographic features in the emergent setting.

**Methods:** We conducted a retrospective chart review of middle cerebral artery (MCA) occlusions treated with MT and confirmed as ICAD or non-ICAD. We recorded common risk factors and radiographic features from CT angiography to identify significant differences between groups. These factors were used in a multivariable logistic regression to create the algorithm. The ROME score was then tested against the ABC^2^D algorithm for predicting ICAD LVO in a prospective cohort.

**Results:** The analysis included 33 ICAD and 327 non-ICAD LVO strokes. ICAD LVO patients were less likely to have atrial fibrillation (9.1% vs 53.8%; [points: 4]) or systolic heart failure with EF≤35% (9.1% vs 27.8%; [points: 1) and more likely to present with progressive or fluctuating symptoms (21.2% vs 4.6%; [points: 1). ICAD patients had a higher incidence of multi-vessel atherosclerotic disease (84.8% vs 37%; [points: 1]), tapered appearance of occlusion (60.6% vs 0.9%; [points: 6]), and extra-cranial ICA atherosclerotic plaque with high-risk features (plaques with lengths ≥1cm or thickness >3mm perpendicular to the long axis of the artery with associated ulceration or with soft plaque component (87.9% vs 37.6%; [points: 4]). AUC for the ROME score was 0.9666 with the highest sensitivity (97%) and specificity (88%) at a cut-off of 9. In the prospective cohort of 201 patients, the ROME score showed 81.3% sensitivity and 98.8% specificity, while the ABC^2^D score showed 90.6% sensitivity and 50.3% specificity.

**Conclusion:** Our scoring system effectively differentiates between ICAD and non-ICAD LVO, with greater specificity than the ABC^2^D score. Future steps will include validation in external databases and clinical trials.

## Introduction

Mechanical thrombectomy (MT) devices were fundamentally designed for the treatment of embolic large vessel occlusions (eLVO), however 10-20% of LVO is caused by large artery atherosclerosis which may not respond comparably in terms of recanalization efficacy and clinical outcomes.^1^ Furthermore, failed thrombectomy is associated with poor neurological outcomes. There is emerging evidence which may suggest that rescue therapy with combination stenting, balloon angioplasty, and intra-arterial glycoprotein IIb/IIIa inhibitors may be effective in revascularization of LVOs due to underlying intracranial atherosclerotic stenosis including those that fail standard thrombectomy.^2,3^ It is also clear that more rapid revascularization with fewer passes is associated with improved neurological outcomes.^4^ Therefore, the differentiation between intracranial ICAD and non-ICAD LVOs as the cause of acute ischemic stroke (AIS) may be important for selecting the ideal MT technique and device(s). Currently, only one prior study by Liao et al. (2022) proposed a new scoring system (ABC^2^D) for reliable differentiation between ICAD and non-ICAD LVOs.^5^ However, the ABC^2^D algorithm has not yet been tested in a prospective study for validation.

The aim of this study was to construct an algorithm for use in the emergent setting to identify ICAD LVOs using clinical risk factors and radiographic features of the LVO. The risk factors for large artery atherosclerosis and cardio-embolism are well described. The former accounts for 10-30% of all LVOs and is more prevalent in African and Asian Americans, those with hypertension, diabetes mellitus (DM), hyperlipidemia, chronic kidney disease (CKD) or end stage renal disease (ESRD), and smoking.^6^ Conversely, the latter is more commonly seen in patients with atrial fibrillation/flutter, recent myocardial infarction, a history of prior embolism, systolic heart failure, patent foramen ovale (PFO), aortic arch atheroma, prosthetic heart valves, and infective endocarditis.^7^ The clinical presentation of patients with underlying ICAD may also differ from the typical eLVO in that the former may present with recent ipsilateral transient ischemic attacks (TIA) and with fluctuating or slowly progressive neurological symptoms, while the latter often presents with abrupt onset of severe deficits.^8^ In addition to the above clinical features, patients with ICAD may be more likely to have radiographic features of atherosclerosis, such as calcifications within the occluded vessel, vessel wall ulceration, and multi-vessel stenosis.^9–11^ The appearance of the occlusion on computed tomography angiography (CTA) and digital subtraction angiography (DSA) may also differ : tapered-appearing occlusions with concomitant Willisian and leptomeningeal collaterals are more commonly observed in ICAD LVOs whereas an abrupt vessel cutoff or a meniscus sign is classically seen in eLVOs.^12, 13^ Several studies have previously described radiographic findings of LVOs that are suggestive of either an embolic thrombus or intrinsic atherosclerotic plaque. Notably, the presence of extra-cranial internal carotid artery (ICA) calcification can significantly increase the risk of LVOs secondary to ICAD, particularly if there is an accompanying stenosis at the bulb of moderate and severe degree per the North American Symptomatic Carotid Endarterectomy Trial (NASCET) criteria, or extra-cranial ICA atherosclerosis with high-risk plaque morphology (i.e. plaques with lengths ≥1cm, thickness >3mm perpendicular to the long axis of the artery with associated ulceration or with soft plaque component).^14–17^ Using the above clinical and radiographic features we have developed a scoring system using retrospective data from all LVOs treated at our institution over the past 7 years to differentiate ICAD and non-ICAD LVOs in the emergent setting. We further tested the ROME score in a prospective cohort trial and compared its reliability to the previously developed ABC^2^D score.

## Methods

We completed a single-center retrospective chart review of all LVO cases presenting in the acute setting that underwent MT between January 2016 to January 2023. The study was approved by the Institutional Review Board and no informed patient consent was required. From the initial roster of 1,321 patients, we excluded all non-MCA trunk (M1) occlusions, including tandem occlusions, patients with incomplete or missing CTA of the head and neck, and patients who did not undergo MT. The final patient roster included 360 patients. The etiology of ischemic stroke in each patient case was reported as eLVO, cryptogenic, or ICAD after thorough clinical investigation during the inpatient admission (Figure 1). Embolic LVO and cryptogenic stroke patients were combined into the non-ICAD category for simplicity of analysis since the artery at the site of the LVO is presumed to be normal. All patients were treated by board certified vascular neurologists and underwent thorough stroke evaluations that included: CT head and CTA in all patients with magnetic resonance imaging (MRI), carotid duplex ultrasound, transcranial Doppler ultrasound, transthoracic echocardiography, transesophageal echocardiography, gated cardiac CTA of the chest, and hypercoagulable panel testing depending on the clinical presentation. All patients had continuous electrocardiographic (ECG) monitoring while hospitalized. Fourteen-day or longer ECG telemetry (external or implanted) was performed in all patients without an identified LVO etiology at discharge.

**Figure 1.**
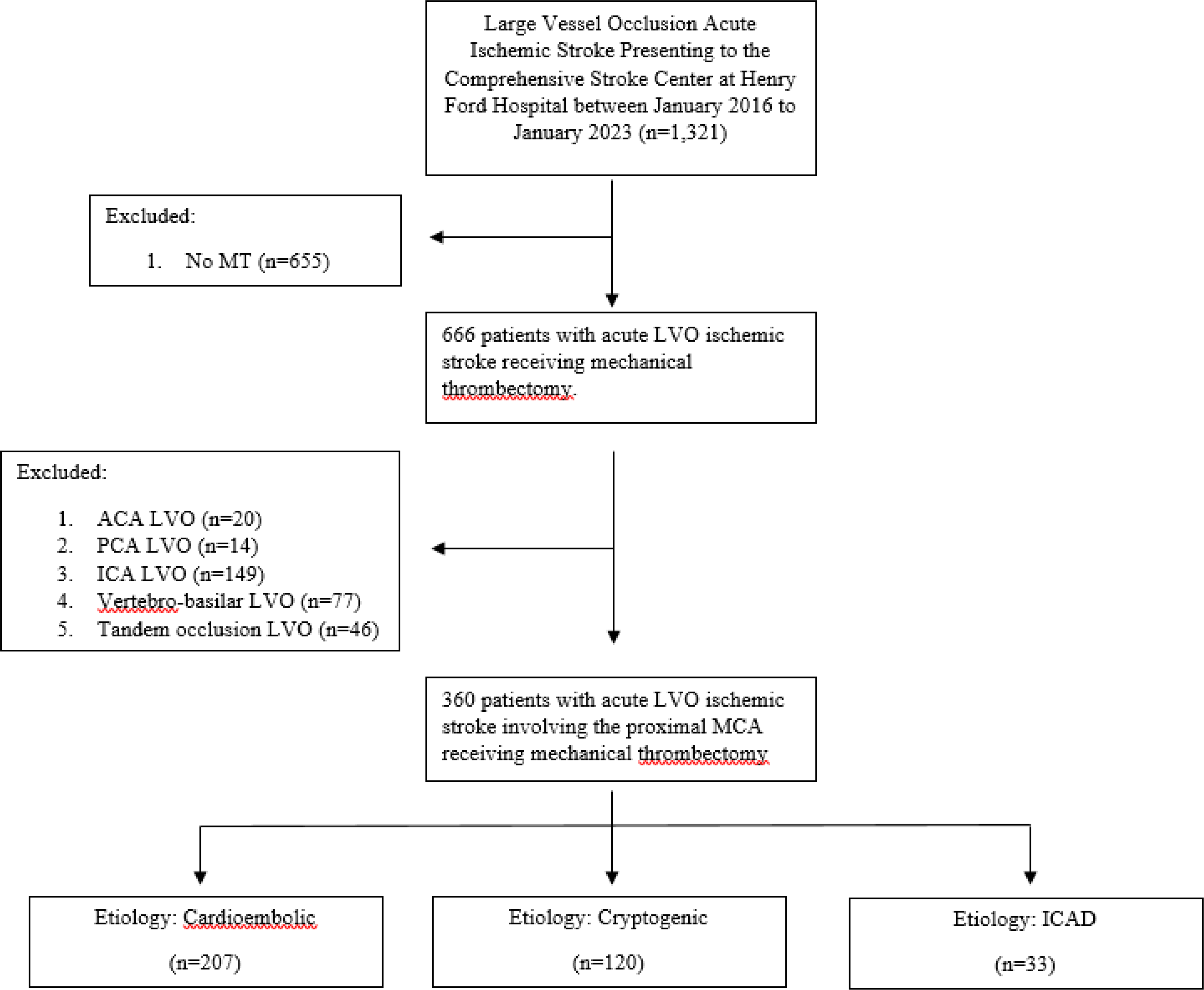
Patient selection criteria for study analysis.

We collected demographic and clinical characteristics as well as the high-risk clinical factors for eLVO and ICAD stroke as documented at the time of stroke admission. eLVO risk factors were determined to be atrial fibrillation (paroxysmal or chronic), anterior wall myocardial infarction within the last 4 weeks, known hypercoagulable states, evidence of systemic emboli or aortic arch atheroma, systolic heart failure with an ejection fraction <35%, and PFO with high risk anatomic features (i.e. PFO ≥3mm in diameter, the presence of atrial septal aneurysm, or hypermobility of the septum during Valsalva maneuver causing a large PFO). ICAD risk factors were determined to be a history of prior ischemic stroke or TIA involving the same vascular territory, African American or Asian American race (risk for intracranial atherosclerosis), history of DM, CKD or ESRD, hypertension, hyperlipidemia, smoking, and progressive or fluctuating stroke symptoms on presentation.

Radiographic findings using the initial CTA head and neck imaging on patient arrival were reviewed by authors of this study and confirmed by formal Neuro-radiology report for the following features: the presence of moderate or severe ICA stenosis at the bifurcation per NASCET criteria, dense vascular calcification of the occluded vessel, multi-vessel intracranial atherosclerotic disease [Figure 2A-F], extra-cranial ICA atherosclerosis with high-risk plaque morphology (i.e. plaques with lengths ≥1cm [Figure 3A-B], thickness >3mm perpendicular to the long axis of the artery with associated ulceration [Figure 3C-D], or with soft plaque component [Figure 3E-F]), and the appearance of the occluded vessel (i.e. abrupt cut-off or tapered with circle of Willis and leptomeningeal collaterals [Figure 4A-D]).^16,17^ We corroborated the CTA head finding of abrupt or tapered LVO with the corresponding DSA images prior to thrombectomy. For tapered-appearing LVOs, we measured the degree of symptomatic vessel stenosis using the Warfarin-Aspirin Symptomatic Intracranial Disease (WASID) criteria on DSA prior to thrombectomy noting those with ≥50% degree of stenosis.^18^ We also reported on the angiographic appearance of the LVO during thrombectomy, noting unique features of the clot associated with eLVOs, including: the presence of contrast flow around the thrombus and the meniscus appearance of the thrombus (i.e. meniscus sign).

**Figure 2.**
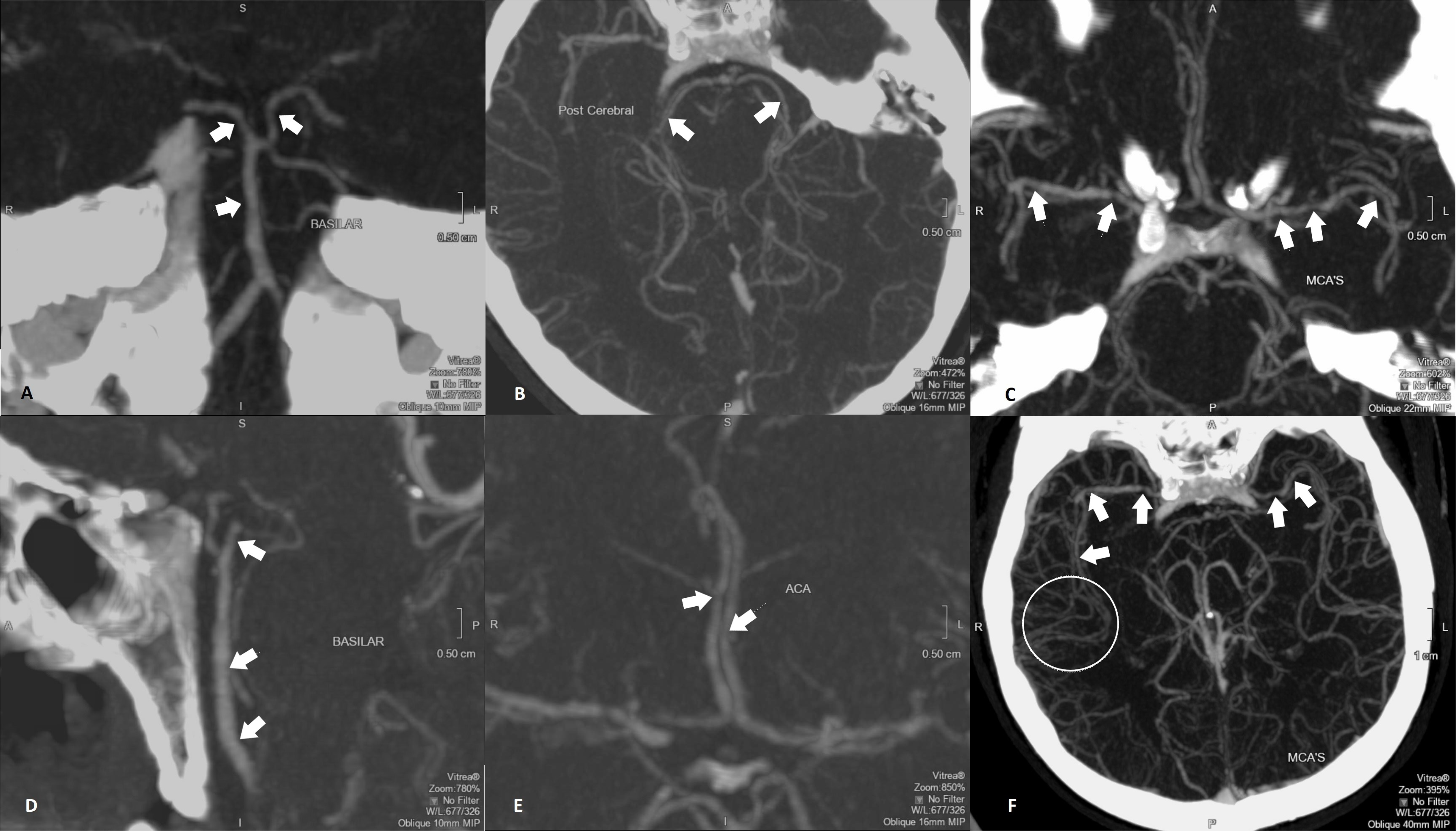
CTA of the head showing intracranial atherosclerotic disease in multiple vessels which appear as a patchy stenosis or beaded pattern indicated by the arrows. *A* – coronal view of the basilar artery. *B –* axial view of bilateral PCAs. *C* – coronal view of the bilateral MCAs. *D* – sagittal view of the basilar artery. *E* – coronal view of the bilateral ACAs. *F* – axial view of the bilateral MCAs with the encircled branches demonstrating Willisian collateralization.

**Figure 3.**
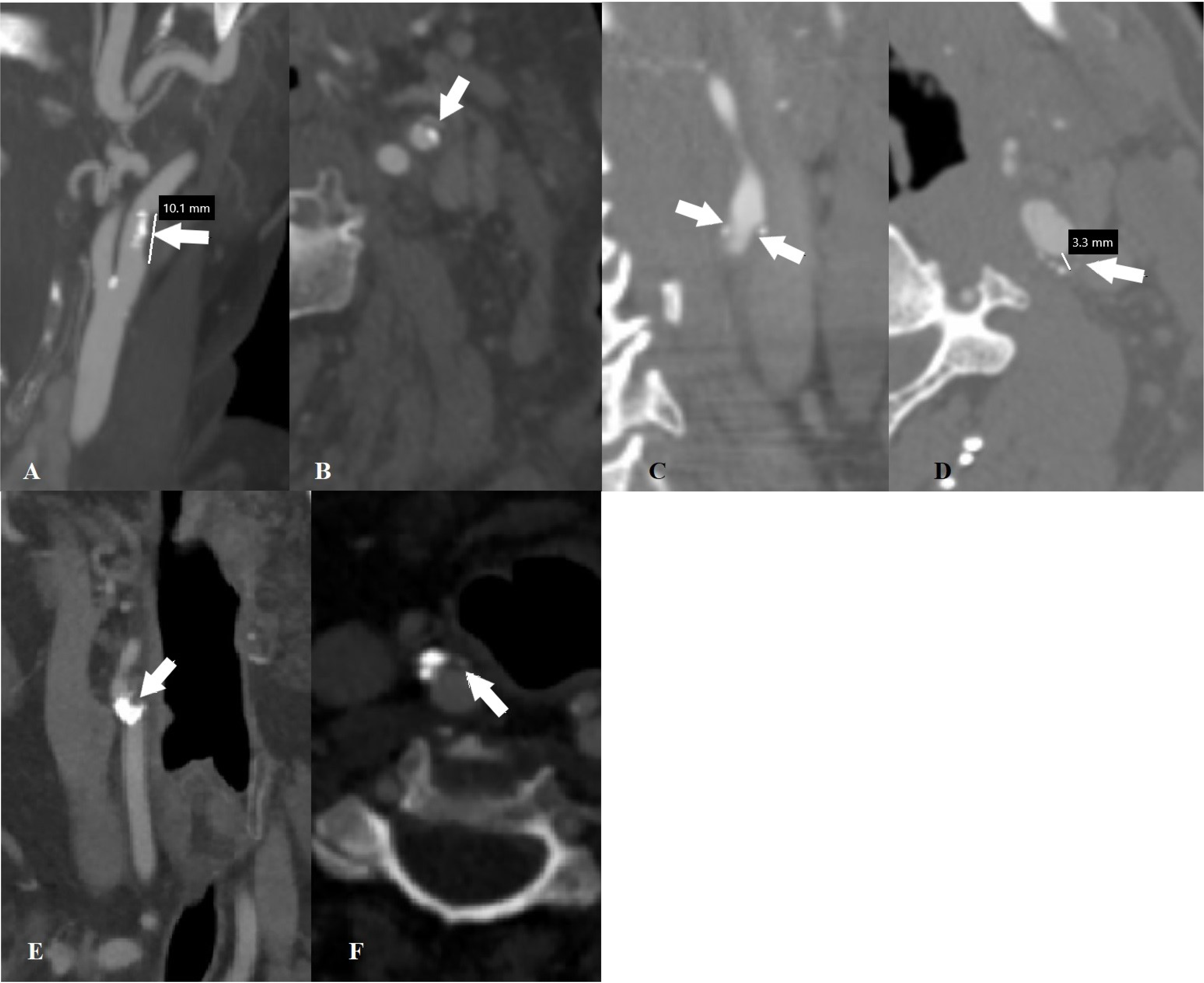
***A-B*** CTA of the neck demonstrating extracranial ICA atherosclerosis with a high-risk soft plaque measuring ≥1cm in longitudinal length. *A* – sagittal view. *B* – axial view. ***C-D*** CTA of the neck demonstrating extracranial ICA atherosclerosis with a high-risk soft plaque with a thickness >3mm perpendicular to the long axis of the artery and ulceration. *A* – sagittal view. *B* – axial view. ***E-F*** CTA of the neck demonstrating extracranial ICA atherosclerosis with a high-risk soft plaque. *A* – sagittal view. *B* – axial view.

**Figure 4.**
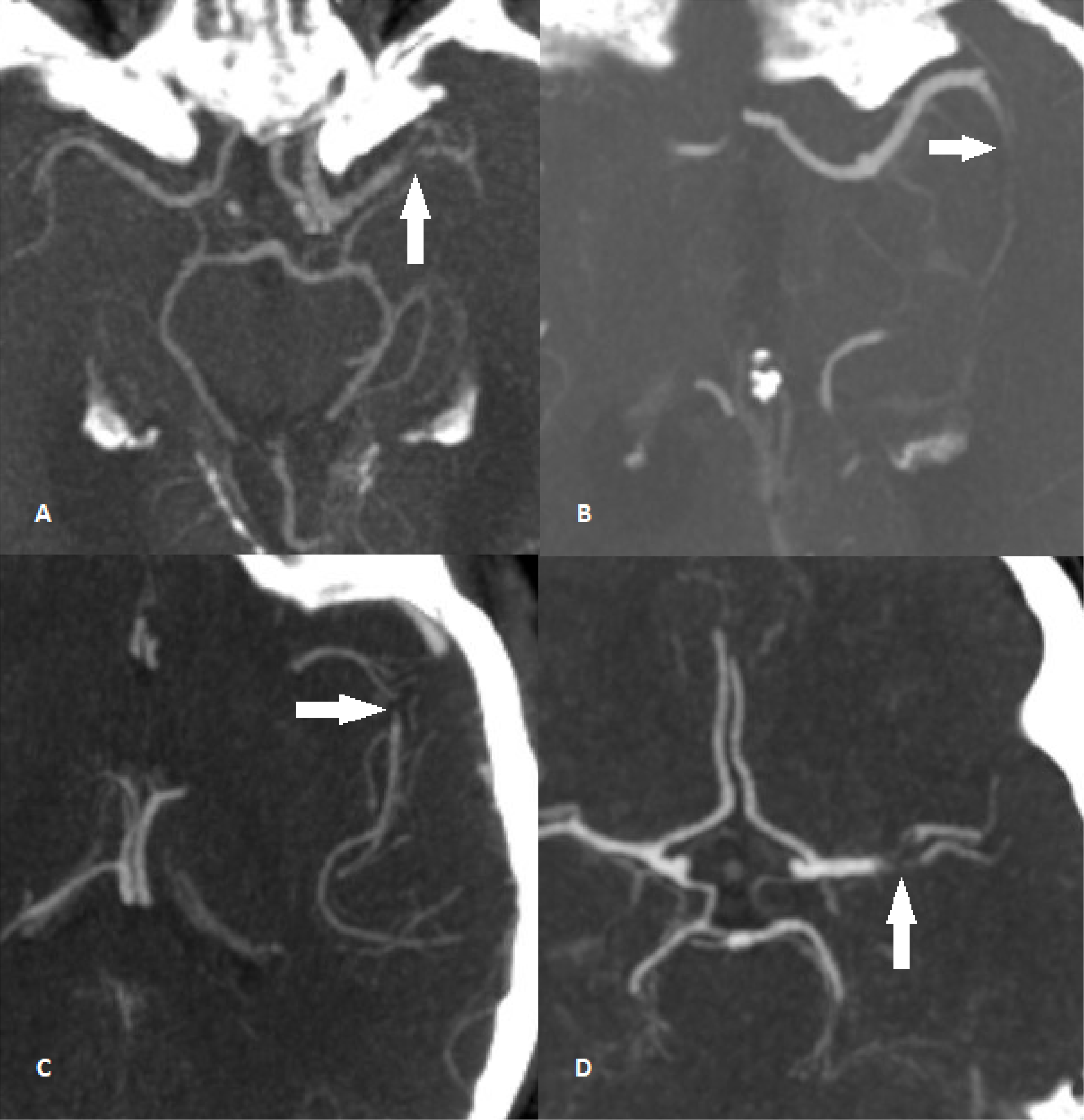
CTA of the head demonstrating intracranial MCA M1 tapered occlusions in four different patients. ***A-C*** *–* axial views. ***D*** – coronal view.

### Prospective Study

We next conducted a single-center prospective cohort study involving all large vessel occlusion (LVO) cases presenting in the acute setting who underwent mechanical thrombectomy (MT) between January 2023 and January 2025. This study was approved by the Institutional Review Board, and informed patient consent was not required. We excluded patients with non-MCA trunk (M1) occlusions, including those with tandem occlusions, as well as patients who were either not candidates for or refused MT. The final cohort included 201 patients.

The ROME score and ABC^2^D score were calculated for each patient on presentation based on known medical history and a review of the initial CTH and CTA head and neck images, assessed by a vascular neurologist and neuroradiologist. ABC^2^D score was calculated based on the following clinical and radiographic features: no prior history of atrial fibrillation (3 points), history of hypertension (1 point), baseline NIHSS score on admission <7 (1 point), absence of the hyperdense vessel sign on CTH (1 point), and history of diabetes mellitus (1 point).^5^ All patients were treated by board-certified vascular neurologists and neuro-interventionalists. Clinical management and further workup were conducted during hospitalization and follow-up clinic visits, which included a comprehensive stroke evaluation to determine the final stroke etiology.

### Statistics

Comparisons among the three groups of patients (ICAD, eLVO, and cryptogenic LVOs) for differences in demographic, clinical risk factors, and imaging characteristics were done using Fisher’s Exact test for categorical variables and Kruskal-Wallis tests for numeric variables. The same analyses were performed comparing the ICAD group to the non-ICAD group (combination of eLVO and cryptogenic patients). To develop our algorithm, we performed a multivariable logistic regression analysis using the significant risk factors and features from the univariate analyses comparing the ICAD and non-ICAD groups. The area under the receiver operating characteristic (ROC) curve (AUC) was computed from the logistic regression model to assess how well the model distinguished between ICAD and non-ICAD groups. The parameter estimates or coefficients for these variables rounded to the nearest integer value were used to determine an appropriate assigned weighted score. Using the sum of these estimates, the sensitivity and specificity were computed for the different score values and the value with that maximized the sum of sensitivity and specificity was considered the ideal cutoff point for differentiating between ICAD and non-ICAD LVOs.

Analysis of the follow-up prospective cohort study using both the ROME and ABC^2^D scores were completed to predict stroke etiology as either ICAD or Non-ICAD stroke. Sensitivity and specificity were calculated to assess how well the scoring algorithms worked by looking at comparisons between predicted stroke etiology and discharge etiology. McNemar’s test was used to compare sensitivity and specificity between the ROME score and ABC^2^D score. The significance level was set at 0.05 for all analyses. All analyses were performed using SAS software version 9.4 (SAS Institute Inc, Cary, NC, USA).

## Results

Out of the 360 patients, 207 were due to eLVOs, 120 were cryptogenic, and 33 were ICAD LVOs. In our initial analysis, we identified significant differences amongst all three groups for the following risk factors: atrial fibrillation (*p<0.0001*), recent anterior myocardial infarction (*p=0.0408*), systolic heart failure with EF <35% (*p<0.0001*), PFO with high risk features (*p=0.0199*), progressive or fluctuating symptoms on presentation (*p=0.0009*), intracranial multi-vessel atherosclerotic disease (*p<0.0001*), abrupt or tapered appearance of occlusion (*p<0.0001*), and extra-cranial ICA atherosclerotic plaque with high-risk features (*p<0.0001*). The tapered appearance of the MCA in ICAD LVOs on initial CTA correlated with a significantly greater percentage of ≥50% intracranial vessel stenosis per WASID criteria seen on DSA prior to thrombectomy (42.4% in ICAD, 0.0% in CE, and 0.8% in cryptogenic; *p<0.0001*). There was no significant difference amongst groups in DSA findings of contrast flow around the thrombus or the presence of a meniscus sign.

We re-analyzed the data using a composite total of cryptogenic and eLVO stroke patients (non-ICAD) against ICAD stroke patients. Patients presenting with ICAD LVOs were significantly less likely to have atrial fibrillation (9.1% vs 53.8%; *p<0.0001*) or systolic heart failure with EF<35% (9.1% vs 27.8%; *p=0.0208*) and more likely to present with progressive or fluctuating symptoms (21.2% vs 4.6%; *p=0.0018*), intracranial multi-vessel atherosclerotic disease (84.8% vs 37%; *p<0.0001*), tapered appearance of occlusion with associated Willisian and leptomeningeal collaterals (60.6% vs 0.9%; *p<0.0001*), and extra-cranial ICA atherosclerotic plaque with high-risk features (87.9% vs 37.6%; *p<0.0001*) (Table 1).

**Table 1.** Compilation of data categorized into the ICAD LVO strokes and non-ICAD LVO strokes (i.e. cryptogenic and eLVO) with corresponding total number of patients and percentages. There were 33 ICAD LVO strokes and 327 non-ICAD LVO strokes. There were significant differences between groups for history of atrial fibrillation, systolic heart failure with EF<35%, and progressive or fluctuating symptoms on presentation, as well as on radiographic imaging findings of moderate or severe MCA stenosis (per WASID criteria), multi-vessel intracranial atherosclerotic disease, abrupt or tapered appearance of occlusion, and extra-cranial ICA atherosclerosis with high risk plaque features (*p<0.05*).

|  | **Non-ICAD**<br>**(N=327)** | **ICAD**<br>**(N=33)** | **Total**<br>**(N=360)** | **P-value** |
| --- | --- | --- | --- | --- |
| **Race, n (%)** |  |  |  | 0.6420<sup>2</sup> |
| African American | 123 (37.6%) | 10 (30.3%) | 133 (36.9%) |  |
| Asian | 19 (5.8%) | 1 (3.0%) | 20 (5.6%) |  |
| White | 185 (56.6%) | 22 (66.7%) | 207 (57.5%) |  |
| **Atrial Fibrillation, n (%)** |  |  |  | <.0001<sup>2</sup> |
| No | 151 (46.2%) | 30 (90.9%) | 181 (50.3%) |  |
| Yes | 176 (53.8%) | 3 (9.1%) | 179 (49.7%) |  |
| **Recent MI, n (%)** |  |  |  | 0.3842<sup>2</sup> |
| No | 310 (94.8%) | 33 (100.0%) | 343 (95.3%) |  |
| Yes | 17 (5.2%) | 0 (0.0%) | 17 (4.7%) |  |
| **Hypercoagulable State, n (%)** |  |  |  | 0.3277<sup>2</sup> |
| No | 269 (82.3%) | 30 (90.9%) | 299 (83.1%) |  |
| Yes | 58 (17.7%) | 3 (9.1%) | 61 (16.9%) |  |
| **Systemic Emboli, n (%)** |  |  |  | 0.2275<sup>2</sup> |
| No | 292 (89.3%) | 32 (97.0%) | 324 (90.0%) |  |
| Yes | 35 (10.7%) | 1 (3.0%) | 36 (10.0%) |  |
| **Systolic Heart Failure with EF**<br>**<35% , n (%)** |  |  |  | 0.0208<sup>2</sup> |
| No | 236 (72.2%) | 30 (90.9%) | 266 (73.9%) |  |
| Yes | 91 (27.8%) | 3 (9.1%) | 94 (26.1%) |  |
| **PFO with High-Risk Features,**<br>**n (%)** |  |  |  | 1.0000<sup>2</sup> |
| No | 309 (94.5%) | 32 (97.0%) | 341 (94.7%) |  |
| Yes | 18 (5.5%) | 1 (3.0%) | 19 (5.3%) |  |
| **Recurrent TIA or Strokes in**<br>**the Same Vascular**<br>**Distribution, n (%)** |  |  |  | 0.6893<sup>2</sup> |
| No | 310 (94.8%) | 31 (93.9%) | 341 (94.7%) |  |
| Yes | 17 (5.2%) | 2 (6.1%) | 19 (5.3%) |  |
| **DM, n (%)** |  |  |  | 0.8467<sup>2</sup> |
| No | 223 (68.2%) | 22 (66.7%) | 245 (68.1%) |  |
| Yes | 104 (31.8%) | 11 (33.3%) | 115 (31.9%) |  |
| **CKD or ESRD, n (%)** |  |  |  | 0.5563<sup>2</sup> |
| No | 293 (89.6%) | 31 (93.9%) | 324 (90.0%) |  |
| Yes | 34 (10.4%) | 2 (6.1%) | 36 (10.0%) |  |
| **Vascular Risk Factors, n (%)** |  |  |  | 0.7823<sup>2</sup> |
| No | 42 (12.8%) | 3 (9.1%) | 45 (12.5%) |  |
| Yes | 285 (87.2%) | 30 (90.9%) | 315 (87.5%) |  |
| **Progressive or Fluctuating Symptoms, n (%)** |  |  |  | **0.0018<sup>2</sup>** |
| No | 312 (95.4%) | 26 (78.8%) | 338 (93.9%) |  |
| Yes | 15 (4.6%) | 7 (21.2%) | 22 (6.1%) |  |
| **Moderate or Severe Extracranial ICA Stenosis (NASCET Criteria), n (%)** |  |  |  | 0.0712<sup>2</sup> |
| No | 313 (95.7%) | 29 (87.9%) | 342 (95.0%) |  |
| Yes | 14 (4.3%) | 4 (12.1%) | 18 (5.0%) |  |
| **Moderate or Severe MCA Stenosis (WASID Criteria), n (%)** |  |  |  | **<.0001<sup>2</sup>** |
| No | 326 (99.7%) | 19 (57.6%) | 345 (95.8%) |  |
| Yes | 1 (0.3%) | 14 (42.4%) | 15 (4.2%) |  |
| **Dense Vascular Calcification of the Occluded Vessel, n (%)** |  |  |  | 1.0000<sup>2</sup> |
| No | 326 (99.7%) | 33 (100.0%) | 359 (99.7%) |  |
| Yes | 1 (0.3%) | 0 (0.0%) | 1 (0.3%) |  |
| **Multi-vessel Intracranial Atherosclerotic Disease, n (%)** |  |  |  | **<.0001<sup>2</sup>** |
| No | 206 (63.0%) | 5 (15.2%) | 211 (58.6%) |  |
| Yes | 121 (37.0%) | 28 (84.8%) | 149 (41.4%) |  |
| **Abrupt or Tapered Occlusion Appearance, n (%)** |  |  |  | **<.0001<sup>2</sup>** |
| Abrupt | 324 (99.1%) | 13 (39.4%) | 337 (93.6%) |  |
| Tapered | 3 (0.9%) | 20 (60.6%) | 23 (6.4%) |  |
| **Extracranial ICA Atherosclerosis with High-Risk Plaque Morphology, n (%)** |  |  |  | **<.0001<sup>2</sup>** |
| No | 204 (62.4%) | 4 (12.1%) | 208 (57.8%) |  |
| Yes | 123 (37.6%) | 29 (87.9%) | 152 (42.2%) |  |
| **Contrast Flow around the Thrombus, n (%)** |  |  |  | 0.0601<sup>2</sup> |
| No | 260 (79.5%) | 31 (93.9%) | 291 (80.8%) |  |
| Yes | 67 (20.5%) | 2 (6.1%) | 69 (19.2%) |  |
| **Meniscus Sign, n (%)** |  |  |  | 0.3260<sup>2</sup> |
| No | 270 (82.6%) | 30 (90.9%) | 300 (83.3%) |  |
| Yes | 57 (17.4%) | 3 (9.1%) | 60 (16.7%) |  |
| <sup>1</sup>Kruskal-Wallis p-value; <sup>2</sup>Fisher Exact p-value |  |  |  |  |

The score estimates and corresponding odds ratios from the multivariable logistic regression analysis are presented in Table 2. The estimated score value was rounded to the nearest integer value and reflected the likelihood that the LVO is secondary to ICAD. A score of 4 was assigned to patients without a history of atrial fibrillation; a score of 1 was assigned to patients without a history of systolic heart failure with an EF<35%; a score of 1 was assigned to patients presenting with progressive or fluctuating symptoms; a score of 1 was assigned to patients multi-vessel intracranial atherosclerotic vessels; a score of 4 was assigned to patients with extra-cranial ICA stenosis and atherosclerotic plaque with high-risk features; and a score of 6 was assigned to patients with tapered appearing occlusions with associated Willisian and leptomeningeal collaterals. The AUC computed was 0.9666 (95% CI=0.9370, 0.9962) and the highest combination of sensitivity and specificity (97% and 88%, respectively) was associated with a score of 9.

**Table 2.**
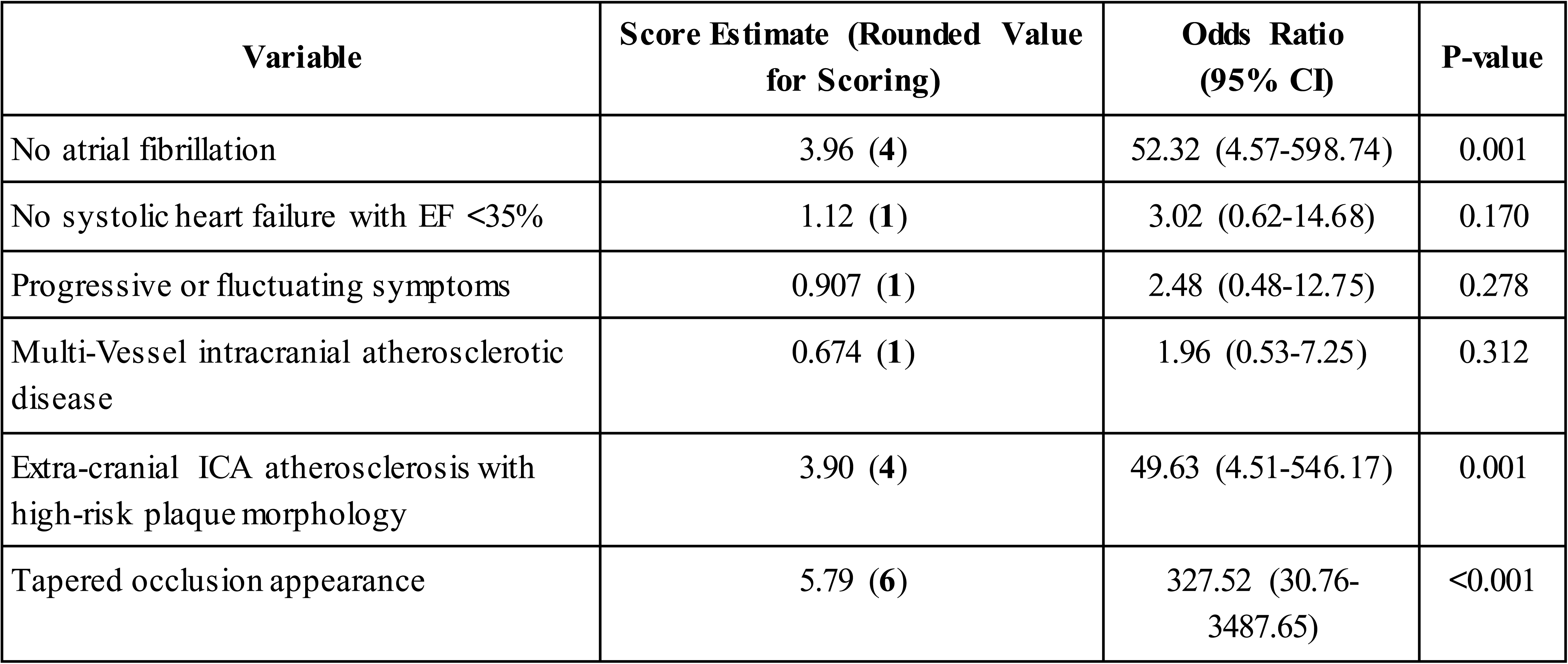
Score estimation using 6 significant clinical and radiographic (i.e. CTA head and neck) variables from Table 2 to discern ICAD from non-ICAD LVOs. The variable, moderate to severe MCA stenosis (per WASID criteria), is excluded given the need for catheter angiography to identify this radiographic feature. A multivariate logistic regression analysis was performed to determine corresponding estimated scores to each variable and the associated odds ratios (95% CI) and p-values.

| Variable | Score Estimate (Rounded Value for Scoring) | Odds Ratio (95% CI) | P-value |
| --- | --- | --- | --- |
| No atrial fibrillation | 3.96 (4) | 52.32 (4.57-598.74) | 0.001 |
| No systolic heart failure with EF <35% | 1.12 (1) | 3.02 (0.62-14.68) | 0.170 |
| Progressive or fluctuating symptoms | 0.907 (1) | 2.48 (0.48-12.75) | 0.278 |
| Multi-Vessel intracranial atherosclerotic disease | 0.674 (1) | 1.96 (0.53-7.25) | 0.312 |
| Extra-cranial ICA atherosclerosis with high-risk plaque morphology | 3.90 (4) | 49.63 (4.51-546.17) | 0.001 |
| Tapered occlusion appearance | 5.79 (6) | 327.52 (30.76-3487.65) | <0.001 |

### Prospective Study

Out of the 201 patients, 169 were non-ICAD strokes (i.e. 96 cardioembolic and 73 cryptogenic) and 32 were ICAD strokes. The ROME score correctly predicted 26 of the 32 ICAD strokes and 167 of the 169 non-ICAD strokes with a sensitivity of 81.3% (95% CI=0.677, 0.948) and specificity of 98.8% (95% CI=0.972, 1.00) (Table 3). The ABC^2^D score correctly predicted 29 of the 32 ICAD strokes and 85 of the 169 non-ICAD strokes with a sensitivity of 90.6% (95% CI=0.805, 1.00) and specificity of 50.3% (95% CI=0.428, 0.578) (Table 4). There was no evidence of a statistically significant difference between the sensitivity of the ROME score and ABC^2^D score (p=0.375). However, the ROME score had a statistically significant higher degree of specificity compared to the ABC^2^D score (p<.0001).

**Table 3.**
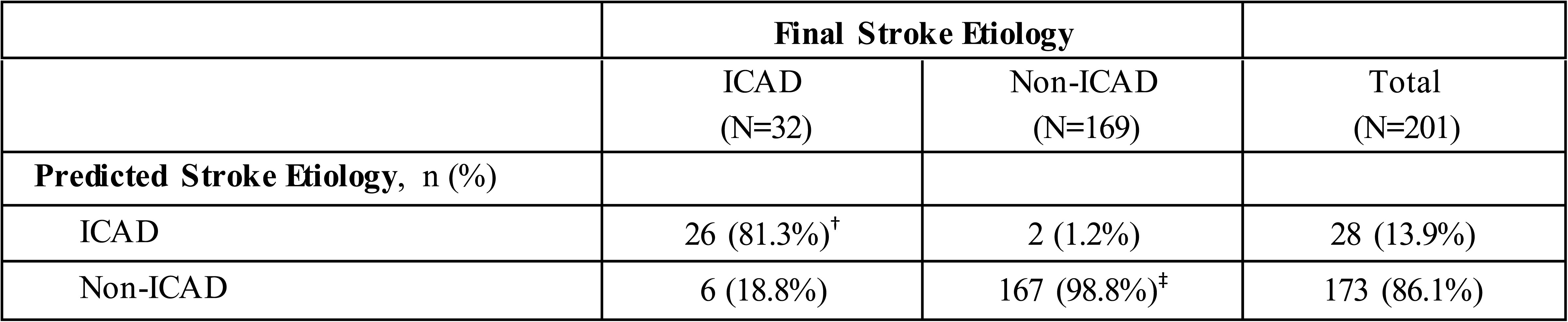
Sensitivity^†^ and specificity^‡^ of the ROME score stroke etiology prediction compared to the final stroke etiology determined at the completion of a comprehensive stroke evaluation during hospitalization and clinic follow-up. Significance level was set at 0.05.

|  | ICAD<br>(N=32) | Non-ICAD<br>(N=169) | Total<br>(N=201) |
| --- | --- | --- | --- |
| **Predicted Stroke Etiology, n (%)** |  |  |  |
| ICAD | 26 (81.3%)<sup>†</sup> | 2 (1.2%) | 28 (13.9%) |
| Non-ICAD | 6 (18.8%) | 167 (98.8%)<sup>‡</sup> | 173 (86.1%) |

**Table 4.** Sensitivity^†^ and specificity^‡^ of the ABC^2^D score stroke etiology prediction compared to the final stroke etiology determined at the completion of a comprehensive stroke evaluation during hospitalization and clinic follow-up. Significance level was set at 0.05.

|  | **Final Stroke Etiology** |  |  |
| --- | --- | --- | --- |
|  | LAA<br>(N=32) | Non-LAA<br>(N=169) | Total<br>(N=201) |
| **Predicted Stroke Etiology, n (%)** |  |  |  |
| LAA | 29 (90.6%)<sup>†</sup> | 84 (49.7%) | 113 (56.2%) |
| Non-LAA | 3 (9.4%) | 85 (50.3%)<sup>‡</sup> | 88 (43.8%) |

## Discussion

The current practice of most neuro-interventionalists when faced with an emergent LVO is to use a stent retriever and/or contact aspiration devices for MT. Although less frequently encountered, MT becomes problematic when faced with ICAD LVOs that are less amenable to either approach and require rescue therapy.^19^ A rapid and easy to compute algorithm in the emergent setting may provide invaluable insight for the neuro-interventionalist to select the optimal tools and techniques to use and reduce the need for device exchanges, reduce procedural costs, and shorten the puncture-to-recanalization time.^20^ Using our devised algorithm, scores <u>>9</u> are indicative of an LVO secondary to ICAD while scores <9 suggest a non-ICAD LVO (i.e. eLVO or cryptogenic) with a sensitivity of 97% and specificity of 88%.

Our prospective cohort study validates the clinical utility of this algorithm with a high degree of sensitivity (81.3%) and specificity (98.8%) for predicting ICAD LVO. When compared to the ABC^2^D score, we report no statistical significance between the sensitivities of the scoring algorithms (p=0.375), however there was a statistically significant difference between the specificities (p<0.0001). These findings suggest that the ROME score is more accurate in identifying non-ICAD LVO in the emergent setting and may therefore be of stronger clinical utility. We postulate several reasons for this significant difference in specificity. First, we find the substantial weighting of the score given to previously known history of atrial fibrillation in the ABC^2^D score drastically impacts the predicted etiology. In our prospective cohort, 26 of the 96 patients presented with a cardioembolic stroke without a known prior history of atrial fibrillation. A majority of these patients were found to have atrial fibrillation during admission or on follow-up. We also suspect that the inherent geographical and lifestyle differences between the populations under study may be contributory, e.g. there is a higher incidence of cardioembolic strokes (25.7% versus 15.8%) and lower incidence of ICAD strokes (14.7% versus 25.4%) in North American populations compared to the Chinese population.^21^ Lastly, while the use of the NIHSS can be helpful in the differentiation of ICAD and CE strokes, it is prone to error in reporting based on subjective evaluation, is limited in posterior circulation strokes, and may be falsely elevated due to previous baseline neurological deficits.

The primary issue faced with ICAD LVOs are the *in-situ* occlusions due to the intrinsic atheroma of the artery. This can result in failure to recanalize as has been reported in several series.^1,22^ In a retrospective study of 1,000 patients, intracranial stenosis was responsible for 70% of failed recanalization of an intracranial occlusion.^1^ Another obstacle with intracranial atherosclerotic occlusions is a much higher rate of re-occlusion of the target vessel. In one meta-analysis, the re-occlusion rate post-MT in ICAD LVOs was 37% as compared to the 2.7% in non-ICAD LVOs with an odds ratio of 23.7 (95% CI, 6.96–80.7).^23^ The above may be overcome with percutaneous transluminal angioplasty and stenting (PTAS) after stentriever or contact aspiration attempts and may result in higher recanalization rates and lower risks of re-occlusion.^24^ Moreover, the negative correlation between procedure duration and neurological outcomes suggests that a direct angioplasty and stenting approach may result in better outcomes in select patient cases. The authors and others have previously published good rates of technical and clinical success with endovascular therapy tailored to the presumed underlying cause of the LVO (i.e. thrombectomy for presumed eLVO and angioplasty and stenting for ICAD LVO). However, this experience predated the modern era of MT with stent-retrievers and contact aspiration catheters.^25,26^

Recently, the Balloon Angioplasty vs Medical Management for Intracranial Artery Stenosis (BASIS) trial has shown promising results in the use of rescue angioplasty. This study demonstrated a significantly lower risk of stroke or death within 30 days up to 12 months with balloon angioplasty and medical management compared to medical management alone in patients presenting with symptomatic severe ICAD.^27^ Randomized control trials investigating application of both angioplasty and stenting are still needed. However, several retrospective studies have shown promising results. One analysis by Stracke et al. (2020) reported on findings from 7 stroke centers and found good functional outcomes (i.e. mRS of 0-2) at 90 days in 44.8% of patients presenting with either an anterior or posterior LVO who received rescue PTAS.^28^ The Stenting and Angioplasty in Neurothrombectomy (SAINT) study was a retrospective analysis of prospective data collected from 14 participating stroke centers. This study evaluated outcomes in patients presenting with an LVO of the anterior circulation who underwent PTAS after failed recanalization. The results demonstrated a significant improvement in the mRS distribution, rates of functional independence (34.6% versus 6.5%), and lower mortality rates at 90 days (29.9% versus 43%) in patients undergone PTAS.^29^ Other retrospective studies have demonstrated improved rates of arterial recanalization (74-98%), low rates of symptomatic intracerebral hemorrhage (0-13%), improved functional outcomes at 3 months (mRS of 0-2 between 34-80%), and lower vessel re-occlusion rates in patients undergone PTAS.^30,31^ Moreover, a few radiographic findings may indicate the need for rescue therapy, including absence of or limited collaterals or a large penumbra with smaller infarct cores.^10^ The results of the ongoing Registry of Emergent Large Vessel Occlusion Due to Intracranial Stenosis (RESCUE-ICAS) trial will also shed more light on the outcomes of ICAD-LVO patients managed medically versus rescue angioplasty and stenting.^32^

Our approach for constructing this algorithm was contingent upon identifying the most commonly encountered and strongly affiliated risk factors for eLVOs and ICAD LVOs. Our dataset identified 9% of the patients as having ICAD LVOs which is consistent with what is reported in the literature.^6^ We excluded several cardio-embolic risk factors which are infrequently encountered in the general patient population presenting with LVO ischemic strokes, including: thrombophilia, prosthetic heart valves, infective endocarditis, papillary fibroelastoma, myxoma, and mitral calcification. Instead, we focused on clinical characteristics that are frequently encountered and can be identified through historical account or basic tests during the acute phase of a presenting LVO, such as: atrial fibrillation (paroxysmal or chronic), systolic heart failure with an ejection fraction ≤35%, recent myocardial infarction (≤4 weeks), history of known PFO with high-risk features (i.e. atrial septal aneurysm, atrial hypermobility, or PFO sizes ≥2mm), known hypercoagulability, history of systemic emboli or aortic arch atheroma, prior ischemic stroke or TIA in the same vascular territory, African American or Asian race, history of DM, CKD or ESRD, vascular risk factors (i.e. hypertension, hyperlipidemia, and smoking history), and whether the presenting stroke symptoms were progressive or fluctuating in nature.

Our initial analysis revealed an overlap in the proportion of clinical risk factors present in patients with eLVOs and cryptogenic LVOs, except for history of known PFO and recent anterior MI. This finding suggests that patients with cryptogenic strokes may have experienced a cardio-embolic event for which a source remains unidentified. This is consistent with literature reports and perspectives – a phenomenon commonly referred to as embolic stroke of undetermined source (ESUS) that is often revealed later as cardio-embolic on further workup.^33^ Therefore, patients diagnosed with a stroke of undetermined etiology may indeed reflect the embolic phenomenon of LVO infarction that is inherently identical to the pathogenic mechanism of cardio-embolic LVOs – i.e. an embolic thrombus which may be of cardiac or arterial source. Hence, under this premise we can differentiate ICAD from non-ICAD LVOs.

Unexpectedly, several risk factors were not significantly different between ICAD and non-ICAD groups, including hypercoagulability, history of systemic emboli or aortic arch atheroma, PFO with high-risk features, recurrent strokes or TIA in the same vascular territory, DM, CKD or ESRD, and vascular risk factors (i.e. hypertension, hyperlipidemia, and smoking history). For instance, approximately 90% of all patients in both groups had multiple vascular risk factors and DM. Therefore, the combined use of both clinical and CTA head and neck features were necessary in constructing an algorithm. Using a multivariable logistic regression model, we identified atrial fibrillation, systolic heart failure with an EF<35%, radiographic findings of intracranial multi-vessel atherosclerotic disease, extra-cranial ICA atherosclerotic plaque disease with high-risk features, and tapered appearance of the LVO with Willisian and leptomeningeal collaterals as significantly different between ICAD and non-ICAD LVOs such that a scoring system could be effectively created with a high sensitivity and specificity for predicting ICAD LVOs.

We also studied whether the use of DSA imaging findings could provide additional insight into the etiology of the LVO in conjunction to the clinical and CTA criteria in the algorithm. For example, there is a well-established correlation between non-tapered occlusions, such as the meniscus-appearing clot with or without surrounding contrast flow, and cardio-embolic thrombi that can be appreciated on both DSA and CTA studies.^12,34^ However, when assessing this in our cohort, we did not find a significant difference in clot appearance between ICAD and non-ICAD LVO groups. There is also a well reported correlation between the location of the occlusion on DSA imaging and the likelihood of atherosclerotic plaque disease and thrombosis. Specifically, branching-site occlusions are more likely to be due to an embolic thrombus while truncal type occlusions are more likely secondary to atherosclerotic plaque disease.^11^ However, the issue faced with this observational correlation is the potential for clot migration wherein a thrombus initially observed in a proximal vessel on CTA is seen more distally on DSA. A notable post-hoc analysis of the Randomized Study of EVT With Versus Without Intravenous Recombinant Tissue-Type Plasminogen Activator in Acute Stroke with ICA and M1 Occlusion (SKIP) trial by Higashida et al. (2021) highlighted that 38% of the patients presenting with a proximal MCA occlusion exhibited clot migration to the distal MCA or first order branches on subsequent DSA, whereas only 9% of patients presenting with an ICA occlusion exhibited clot migration.^35^ Therefore, while clot location on CTA could be potentially helpful in distinguishing between thrombus and atherosclerotic plaque, it may not always present identically on DSA studies particularly when involving the MCA trunk.

Our study has several limitations, including those inherent to a retrospective study. The definitions of stroke etiology were not standardized although all patients were treated by board certified vascular neurologists at a major academic medical center with standardized internal protocols. Another limitation was the relatively lower number of ICAD LVOs in our patient population. Although reflective of the percentage seen in the general population of LVO strokes, including more patients within this cohort may further delineate the presence or absence of significant differences in risk factors. Validation of our algorithm with an external patient database which includes a larger roster of ICAD LVO patients would bolster our findings and support testing in a clinical trial study. We recognize that MCA LVOs represent only a proportion of LVOs encountered by stroke clinicians and interventionalists. A future direction will be to extend this algorithm for use in cervical ICA LVOs and posterior circulation strokes.

In summary our clinical and imaging algorithm can reliably differentiate ICAD LVO from non-ICAD LVO. This algorithm will need to be validated in a larger prospective dataset. The prediction of the etiology of the LVO prior to MT may help guide the neuro-interventional approach and devices.

## Data Availability

Data acquired in study will be available upon individual request.

## Acknowledgements

None

